# Prompt Engineering in Large Language Models for Patient Education: A Systematic Review

**DOI:** 10.1101/2025.03.28.25324834

**Authors:** Aya Mudrik, Girish N Nadkarni, Orly Efros, Shelly Soffer, Eyal Klang

**Affiliations:** Ben-Gurion University of the Negev, Be’er Sheva, Israel; The Windreich Department of Artificial Intelligence and Human Health, Mount Sinai Medical Center, NY, USA; The Charles Bronfman Institute of Personalized Medicine, Icahn School of Medicine at Mount Sinai, New York, NY, USA; National Hemophilia Center and Institute of Thrombosis & Hemostasis, Chaim Sheba Medical Center, Tel Hashomer, Israel; Faculty of Medicine, Tel Aviv University, Tel Aviv, Israel; Institute of Hematology, Davidoff Cancer Center, Rabin Medical Center, Petah-Tikva, Israel

**Keywords:** Prompt Engineering, Large Language Models, Patient Education, Readability, Health Literacy

## Abstract

**Background:** Large language models (LLMs) have shown promise in generating patient-friendly medical content, but their outputs often vary in accuracy, readability, and relevance. Prompt engineering—structuring inputs to guide LLM responses—may improve the quality of educational materials, yet its impact on patient education remains unclear.

**Objectives:** To systematically review whether prompt engineering improves readability, accuracy, and usability of LLM-generated content for patient education.

**Methods:** We conducted a systematic review in accordance with PRISMA guidelines. PubMed, Scopus, and Web of Science were searched for original studies evaluating prompt engineering techniques in patient education. Data were extracted on prompt types, LLM models used, and outcomes. Risk of bias was assessed using the QUADAS-2 tool, and a narrative synthesis was performed.

**Results:** Our search identified five studies that met our criteria, focusing on answering patient questions and generating medical information. Prompt engineering techniques included instruction-based, elaborated, role-defining, scene-defining, and domain-specific prompts. Structured prompting improved accuracy and comprehensiveness in several cases, particularly when specific formats or custom instructions were used. Readability gains were notable when prompts explicitly requested simpler language and reading levels, though some strategies unintentionally increased complexity. Variability in effectiveness across LLMs and prompt types was observed.

**Conclusions:** Prompt engineering can enhance the clarity and, in some cases, the accuracy of LLM-generated patient education materials. However, benefits vary by model and strategy. Standardized approaches and further research are needed to optimize prompts, minimize bias, and support reliable, accessible patient communication.

## Introduction

Informed patients tend to follow treatment plans more reliably, take part in decisions, and achieve better outcomes.^1–3^ Clinicians often provide written materials after visits to help patients remember and understand what was discussed. However, existing resources often exceed patients’ literacy levels, incorporate overly technical language, or contain inaccuracies.^4^

Large language models (LLMs), such as OpenAI’s GPT, show promise for generating patient-friendly medical information.^5,6^ However, their reliability varies considerably, potentially introducing errors that could compromise patient decision-making and adherence.^6–8^

Prompt engineering, the structured refinement of inputs to improve LLM outputs, shows potential for enhancing the quality of patient-directed information.^9^ However, the impact of specific prompt engineering strategies on patient education is not well defined.

We systematically reviewed the literature to see whether prompt engineering improves readability, accuracy, and usability in LLM-generated patient education materials.

### Glossary of AI Terms

AI refers to computational systems that perform tasks requiring human-like intelligence, such as decision-making, pattern recognition, and language understanding. It encompasses various subfields, including machine learning (ML), deep learning (DL) and natural language processing (NLP). ^8,10^

#### Machine Learning (ML)

A form of AI in which systems improve their performance over time by learning from data rather than following explicit programming. ML models process large datasets, identify patterns, and generate predictions based on the insights they derive.^8,11^

#### Natural Language Processing (NLP)

NLP is a branch of AI that enables machines to understand and generate human language. NLP techniques leverage statistical models and DL, particularly transformers, to process text.^12^

#### Deep learning (DL)

A subfield of AI that uses artificial neural networks (ANNs) with multiple layers to recognize patterns and learn from large datasets. These networks mimic biological neural networks. A single artificial “neuron” is akin to a simple logistic regression unit. Multiple “neurons” are connected to create deep layers. ANNs enable complex tasks such as image recognition and speech understanding.^8,13,14^

#### Transformers

Transformers are a class of DL that process sequential data in parallel using a mechanism called self-attention. Unlike previous DL models that rely on recurrence or convolutions, transformers can weigh the importance of all words in a sentence simultaneously, leading to more context-aware language models.^8,15^

#### Large Language Models (LLMs)

LLMs are DL models trained on vast amounts of text data to perform natural language tasks such as text generation, question answering, and summarization. They use transformer architectures to process and generate human-like text.^16^ Common examples of popular LLMs include OpenAI’s GPT, Google’s Gemini (formerly known as Bard), and Perplexity. In **Figure 1**, we present a diagram of the way LLMs work.

**Figure 1.**
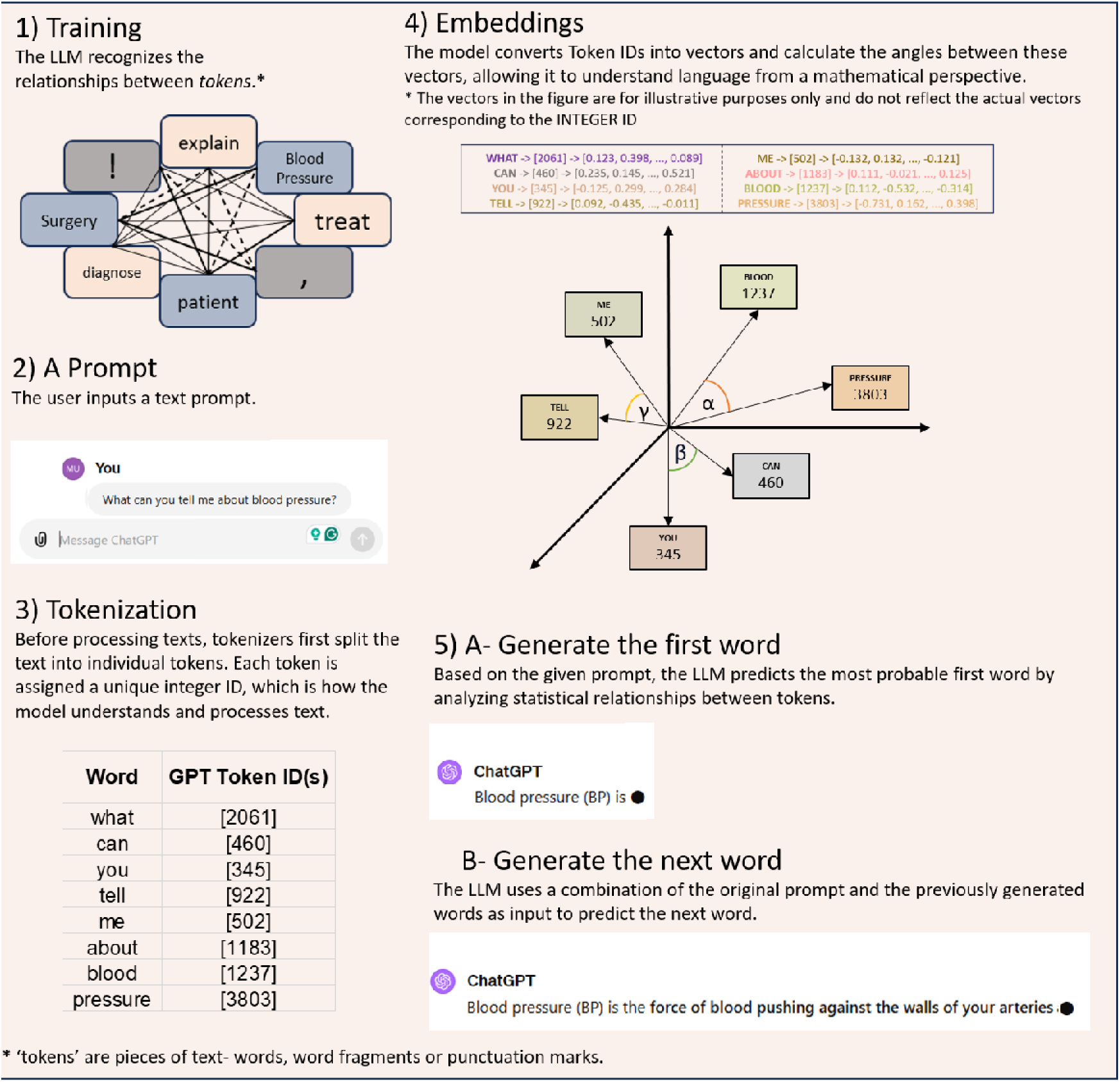
Diagram of the way LLMs work.

#### Prompt Engineering

Prompt engineering is the practice of designing input prompts to optimize the output of LLMs. By structuring prompts effectively, users can guide models to generate more accurate, relevant, and context-aware responses.^17,18^

*Examples of Prompt Engineering Techniques (****Figure 2***):

- *Zero-shot prompt*-Asking the model to complete a task without providing examples or demonstrations.^19^
- *Elaborated prompt-* A refined version of a basic query that includes additional details, clarifications, or constraints.^20^
- *Instruction-based prompt*-Explicitly directing the model on response format and tone.^21^
- *Domain-specific knowledge prompt*-Guiding the model to generate responses using specialized knowledge, terminology, and best practices relevant to a specific field.^22^
- *Role-defining prompt*-Assigning the model a specific identity (such as a physician) to accomplish a task related to that role.^18^
- *Scene-defining prompt*-Simulating a scene related to the addressed task.^18^

**Figure 2.**
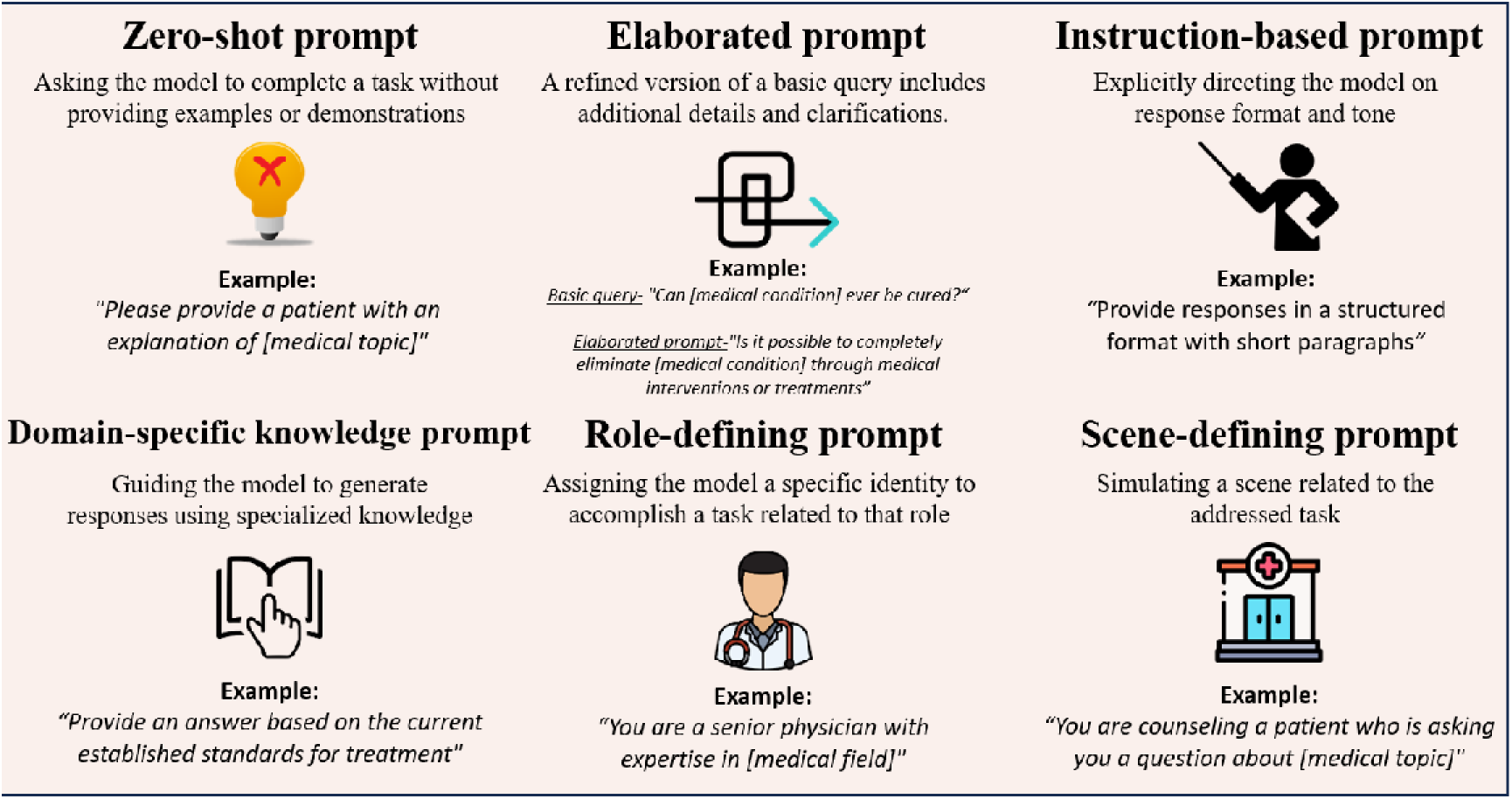
Examples of Prompt Engineering Techniques.

### Readability Metrics for Evaluating LLMs’ Responses

Readability metrics assess the complexity of text and estimate the education level needed for understanding.^23^ The following metrics are commonly used to evaluate LLM-generated responses in medical education:

#### Automated Readability Index (ARI)

Measures readability based on character count per word and words per sentence, providing a U.S. grade level estimate. The ARI score typically ranges from 1 to 14+, where lower scores correspond to elementary-level reading, scores of 7-10 indicate high school-level difficulty, and scores above 11 suggest college-level complexity.^24,25^

#### Flesch-Kincaid Readability Level (FKRL) & Grade Level (FKGL)

FKRL rates readability, while FKGL estimates the school grade level required for understanding based on sentence length and syllables per word. A score of 8-10 is considered accessible to the general public, with higher scores indicating more advanced reading levels.^26–29^ ***Figure 3*** provides examples of texts written at varying Flesch-Kincaid Grade Levels (FKGL), illustrating differences in complexity and readability.

**Figure 3.**
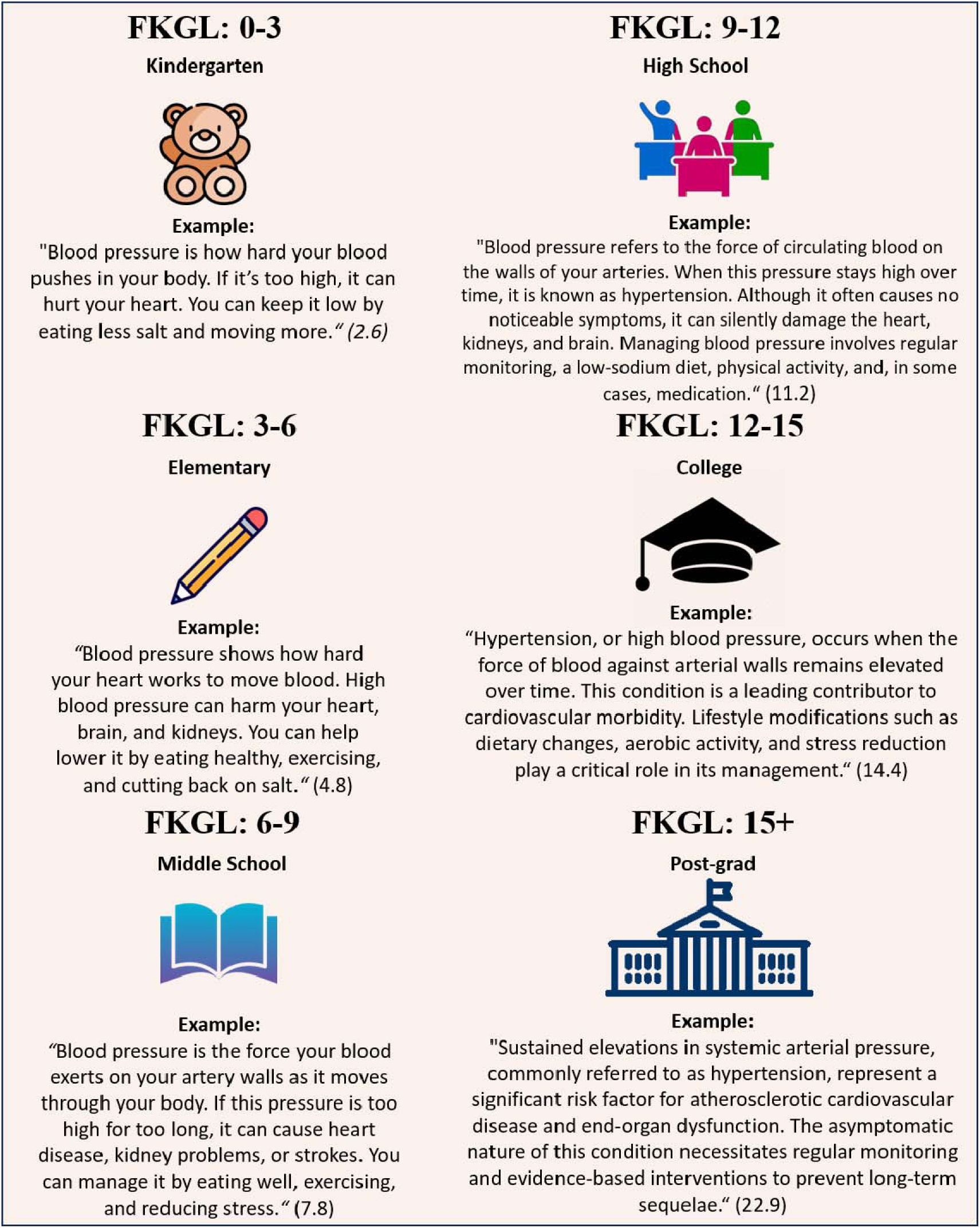
Readability Levels of Patient Education Texts About Blood Pressure: Flesch-Kincaid Grade Level Examples.

#### Simple Measure of Gobbledygook (SMOG)

Estimates education years needed for comprehension by analyzing the number of polysyllabic words. A score of 6-8 is easy to read, while score of 12+ is complex, college-level texts.^27,30^

#### Gunning Fog Index (GFI)

Determines text difficulty using sentence length and complex word frequency, indicating the formal education level required. Scores of 8-10 suggest easy readability, while scores above 12 are typical of academic or technical writing.^31^

#### Coleman-Liau Index (CLI)

Calculates readability based on characters per word and words per sentence, making it well-suited for automated text analysis. Scores of 6-10 are considered readable for most audiences, with higher scores indicating more advanced reading levels.^27,32^ generated patient education materials.

## Methods

### Search Strategy

This systematic review was performed in adherence to the Preferred Reporting Items for Systematic Reviews and Meta-Analyses (PRISMA) guidelines,^33^ incorporating elements from the diagnostic test accuracy extension^34^ and the CHARMS checklist^35^ for reviews of prediction models.

A systematic literature search was executed on February 4, 2025, across PubMed, Web of Science, and Scopus databases. We identified original research that examines the impact of prompt engineering on patient education, particularly in enabling LLMs to generate medical information and answer patient questions.

The search employed specific terms related to prompt engineering within medical databases and was limited to peer-reviewed publications in English. The detailed search strategy is provided in the **Supplementary Materials (’Detailed Search Strategies’)**.

We excluded non-original studies and articles that did not directly address the impact of prompt engineering on patient education. Additionally, references from selected articles were examined to capture any pertinent studies missed in the initial search.The study is registered in the PROSPERO database (CRD420251002182).

### Study Selection

Initial screening of titles and abstracts was conducted independently by two reviewers (AM and SS), with eligibility based on predefined inclusion criteria. Any ambiguities were resolved through full-text assessments. Discrepancies during any stage of the selection process were resolved through consultation with a third reviewer (EK).

### Data Extraction

Data extraction was independently performed by the same two reviewers (AM and SS) using a standardized form designed for this review. Extracted data included publication year, study objectives, types of LLMs utilized, prompting strategies, sample sizes, and primary outcomes.

### Quality Assessment and Risk of Bias

The risk of bias within the evaluated studies was assessed using an adapted version of the Quality Assessment of Diagnostic Accuracy Studies (QUADAS-2) criteria.^36^

### Data Synthesis

Given the heterogeneity in study designs and outcomes, we opted for a narrative synthesis over a meta-analysis. This approach allowed us to summarize the applications, benefits, and challenges associated with the use of prompt engineering in LLMs’ responses to patient medical questions and the generation of medical information, as reported by the included studies.

## Results

A total of 512 articles were retrieved in the initial search **(Supplementary Figure 1)**. Five studies evaluating the impact of prompt engineering on patient education were included.

The characteristics of the studies are presented in **Table 1**. Objectives, reference standard, sample sizes, and main findings are presented in **Table 2**. Prompt formats, examples of prompts, and prompt engineering technique used are presented in **Supplementary Table 1**.

**Table 1.** Characteristics of included studies.

| Category | Title | First Author | Journal/Book | Year |
| --- | --- | --- | --- | --- |
| *Answering Patient Questions* | Prompt engineering with ChatGPT3.5 and GPT4 to improve patient education on retinal diseases | Jung H | Canadian Journal of Ophthalmology | 2024 |
|  | Effects of prompt engineering on large language model performance in response to questions on common ophthalmic conditions | Wu JH | Taiwan Journal of Ophthalmology | 2024 |
|  | Enhancing AI Chatbot Responses in Health Care: The SMART Prompt Structure in Head and Neck Surgery | Vaira LA | OTO Open | 2025 |
| *Generating Medical Information* | De novo generation of colorectal patient educational materials using large language models: Prompt engineering key to improved readability | Ellison IE | Surgery | 2025 |
|  | The Use of Large Language Models to Generate Education Materials about Uveitis | Kianian R | Ophthalmology Retina | 2024 |

**Table 2.**
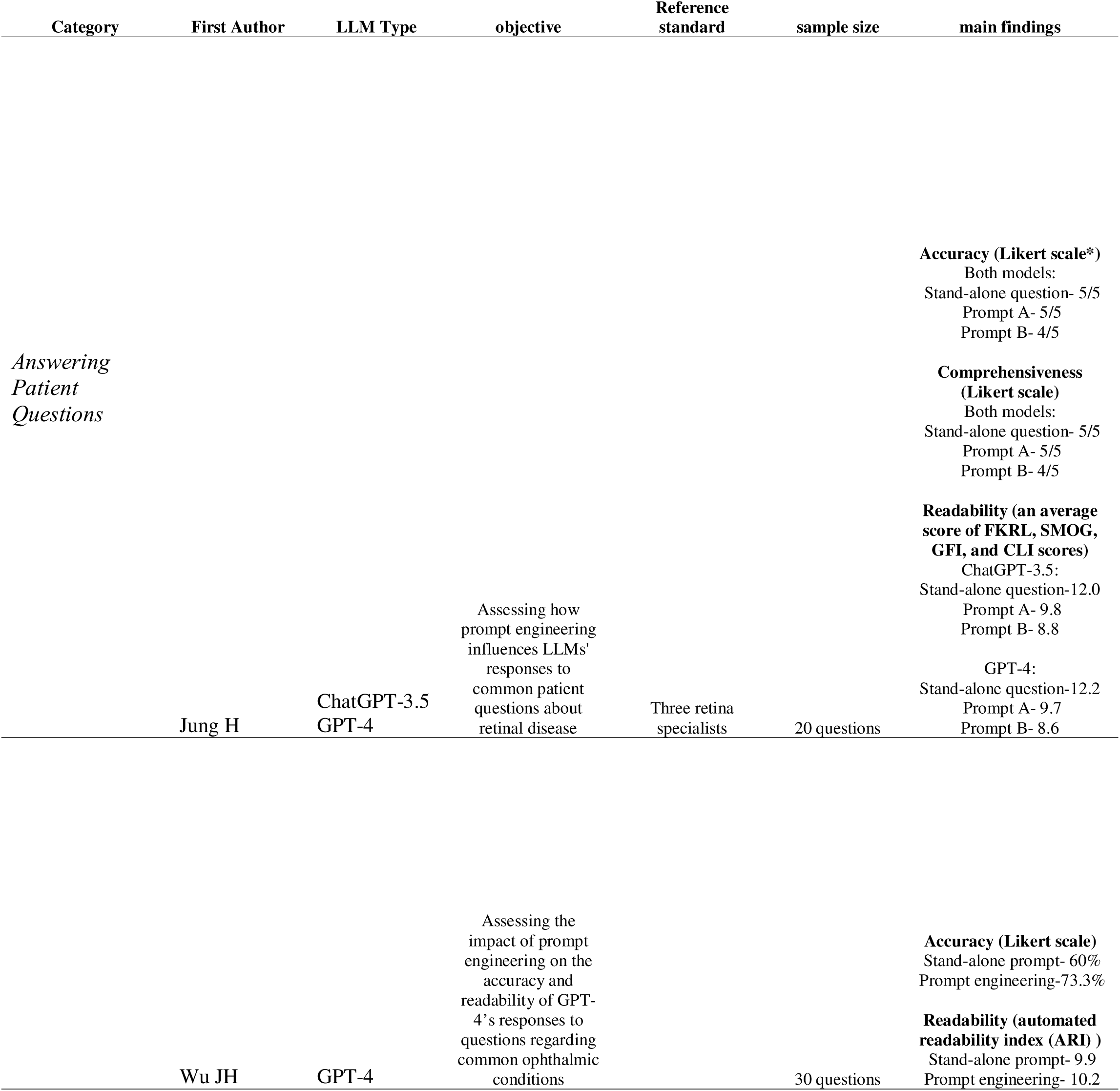
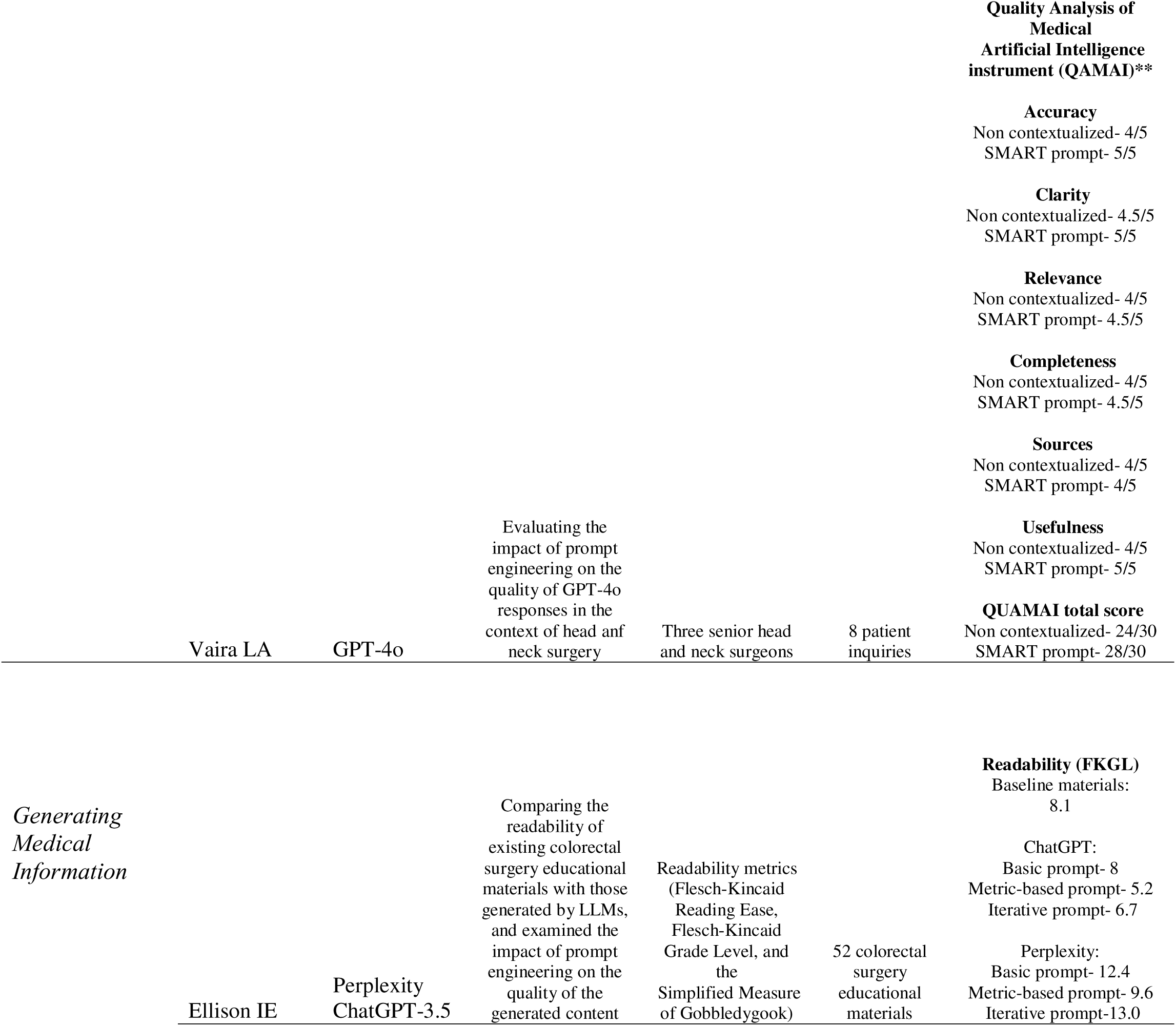
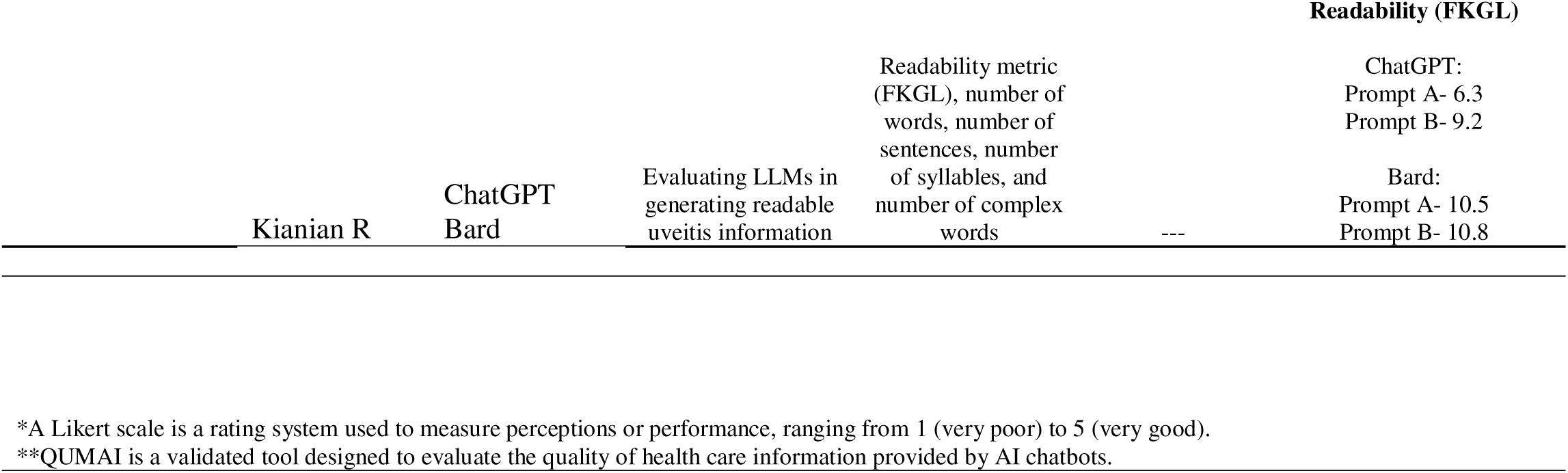
Overview of study designs and key outcomes.

The included studies spanned two categories: answering patient questions and generating medical information. The studies varied in aims, methodologies, and type of prompt engineering used. They also covered a range of medical fields including head and neck surgery, ophthalmology, and general surgery.

Across five studies, researchers tested 12 prompts. All used *zero-shot prompting*, meaning no sample answers were provided. Seven were *instruction-based*, guiding the model to follow a certain tone or format (e.g., simple, clear language). Three were *elaborated prompts*, which built on a basic query with added details. For instance, the *basic prompt* asked if myopia can be cured, while the *elaborated prompt* inquired if medical interventions could fully eliminate it. Three were *scene-defining*, and two were *role-defining*, assigning the model a specific setting or identity. Finally, two prompts included *domain-specific knowledge* to ensure the model used relevant terms and best practices. The different types of prompt engineering are presented in **Figures 2, 4.**

**Figure 4.**
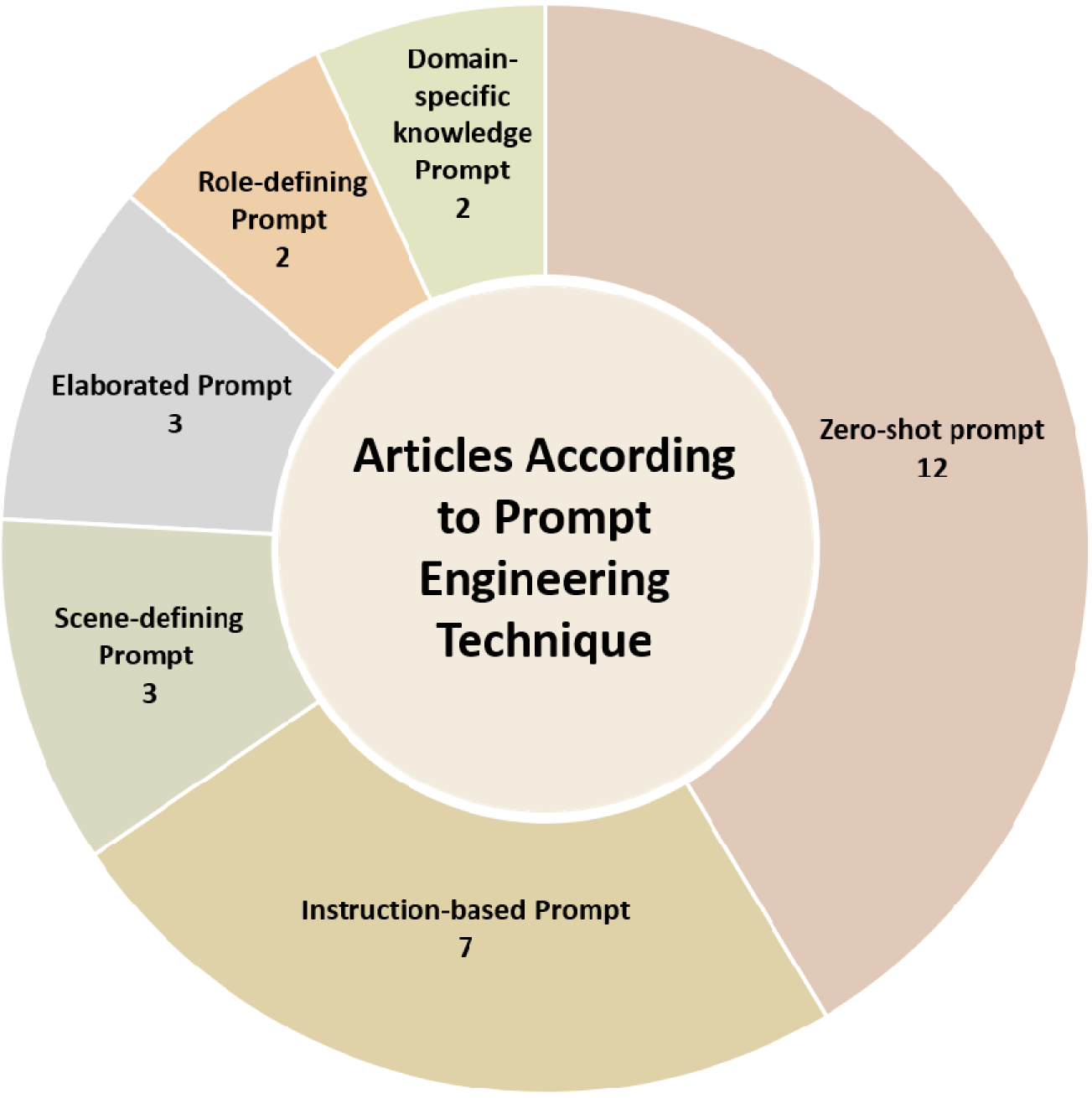
Number of reviewed articles according to Prompt Engineering technique.

The risk of bias and applicability analysis using QUADAS-2 tool is presented in **Supplementary Table 2**.

### Descriptive summary of results

#### Answering Patient Questions

Three studies assessed the impact of prompt engineering on the ability of LLMs to answer patient questions.

Jung H. et al.^27^ studied how prompt engineering affected GPT-3.5 and GPT-4’s responses to 20 common patient questions about retinal disease. They tested three prompt types: a stand-alone prompt, an optimized prompt focused on accuracy, comprehensiveness, and readability (Prompt A), and a shorter version of Prompt A written for an 8th-grade reading level and limited to 300 words (Prompt B) (see **Supplementary Table 1**). Three retina specialists rated the accuracy and comprehensiveness of the responses, while readability was measured using an online tool.

Prompt B led to lower accuracy and comprehensiveness compared to the stand-alone prompt and to Prompt A. However, both Prompts A and B significantly improved readability, with Prompt B producing the most readable responses, using simpler language and fewer technical terms. The presence of empathetic language, disclaimers, and specialist referrals did not vary by prompt type.^29^

Wu JH. et al.^25^ also examined the impact of prompt engineering in ophthalmology. Their study evaluated how prompt engineering affects GPT-4’s accuracy and readability when answering questions about common eye conditions. They collected 30 frequently asked online questions and submitted them to GPT-4, both with and without prompt engineering. To improve responses, they first used GPT-4’s “Custom Instructions” to define the model as the user’s ophthalmologist, aiming for accurate, clear, and professional answers. Then, they used an automated tool called “Prompt Perfect” to rephrase the questions, making them more specific (see **Supplementary Table 1**).

This approach improved accuracy-from 60% with unstructured prompts to 73.3% with prompt-engineered ones. However, the improved prompts led to longer, more complex answers, which made them harder to read.

Another study examined how prompt engineering affects the quality of GPT-4o’s responses in head and neck surgery.^37^ A multidisciplinary team, consisting of two head and neck surgeons, a linguist, and a computer engineer, developed the SMART prompt format. The acronym SMART stands for: **Seeker** (user’s professional identity), **Mission** (purpose of the question), **AI Role** (the role assigned to the AI, such as expert or professor), **Register** (tone and scientific level), and **Targeted Question** (a focused, context-rich query). This format helps guide LLMs to give accurate, relevant answers.

Researchers entered 24 medical questions, including 8 from patients, into GPT-4o, both with and without the SMART format (see **Supplementary Table 1**). Three senior head and neck surgeons evaluated the responses based on accuracy, clarity, relevance, completeness, source quality, and overall usefulness. Responses using the SMART format scored significantly higher in all categories.^40^

#### Generating Medical Information

Two studies assessed the impact of prompt engineering on the ability of LLMs to generate medical information.

Ellison I. et al. compared the readability of existing colorectal surgery educational materials with content generated by LLMs, and examined how prompt engineering affected the quality of the generated text.^29^ Researchers collected 52 educational materials from major academic institutions and tested three types of prompts using Perplexity and GPT-3.5. The **Basic prompt** asked for general patient education materials. The **Iterative prompt** refined this request to a sixth-grade reading level. The **Metric-based prompt** specifically asked for sixth-grade readability, short sentences, and simple words, based on National Institutes of Health (NIH) recommendations (see **Supplementary Table 1**).

Readability scores showed that the original materials were too complex, averaging a 7th to 9th-grade level. Of the LLM-generated texts, only GPT’s Metric-based prompt consistently improved readability. Perplexity’s content was generally harder to read than the original materials, except with the Metric-based prompt, which showed only slight improvement. Iterative prompts had mixed results: GPT-3.5 saw minor gains, while Perplexity performed worse than the original texts.^31^

Kianian R. et al, evaluated how well GPT-3.5 and Bard could generate readable information about uveitis.^28^ The models were given two types of prompts: one asked them to write at a 6th-grade reading level, based on the Flesch-Kincaid Grade Level (FKGL) formula (**Prompt A**), and the other asked them to write information that was “easy to understand” without setting a specific reading level (**Prompt B**) (see **Supplementary Table 1**). The responses were analyzed for FKGL, total words, sentences, syllables, and number of complex words.

For both models, responses from Prompt A had significantly fewer complex words than those from Prompt A. GPT’s Prompt A responses were also easier to understand and had fewer sentences than its Prompt B responses. Bard’s Prompt A responses had fewer words and syllables than Bard’s Prompt B responses. Overall, Prompt B did not perform better than Prompt A in any of the measured areas.^30^

## Discussion

This systematic review explores the effect of prompt engineering on the quality of LLM-generated patient education content. The findings illustrate both the benefits and limitations of structured prompts that aim to enhance readability, accuracy, and relevance in this content. Prompt engineering techniques appear to improve certain aspects of model performance. However, their effectiveness varies, underscoring the need for further refinement and standardization.

### The impact of prompt engineering on LLM performance

The reviewed studies show that prompt engineering—such as role-defining prompts, structured query changes, and readability refinements—can significantly improve LLM responses.

Structured prompts led to increased accuracy and comprehensibility of LLM-generated answers to patient questions.^25,27,37^ For example, ophthalmology-related questions received responses with a higher accuracy score when using prompt engineering compared to stand-alone prompts. However, readability scores varied. Some structured prompts led to more complex language than desired.^25^ This suggests that while structured prompts can improve factual correctness, they may also unintentionally increase text complexity. This can limit accessibility for patients with lower health literacy levels.

Prompt engineering also improved readability of generated content when specific readability constraints were applied.^28,29^ For example, instructing models to generate responses at a sixth-grade reading level, with short sentences and simple wording, resulted in answers with lower reading grade levels. However, when prompts specified only a sixth-grade reading level, without additional constraints on sentence length or word simplicity, results were mixed. GPT-3.5 showed minor readability improvements, whereas Perplexity performed worse than the baseline materials.^29^

### Challenges and limitations of prompt engineering

Despite its advantages, prompt engineering presents challenges. One primary concern is inconsistency across different prompt formats and medical fields.^38^ While some structured prompts enhance accuracy and clarity, others contribute to variability in response quality, particularly when applied across different LLMs. This inconsistency emphasize the need for further research to establish standardized prompt formats.

Another limitation is the potential for prompt engineering to introduce unintended biases.^19^ Given that LLMs generate responses based on their training data, prompts that emphasize specific viewpoints or structures may inadvertently shape the model’s outputs. This is particularly concerning in patient education, where it can affect treatment adherence. Future research should explore methods to ensure that prompt-engineered responses remain unbiased.

Additionally, there is no universally accepted framework for evaluating the success of prompt engineering. While readability metrics such as FKGL and SMOG provide insight into linguistic complexity, they do not fully capture nuances such as engagement and empathy, factors that are equally important in patient education.^39^ Developing standardized evaluation metrics that consider both linguistic and contextual quality will help estimate the effectiveness of prompt engineering in medical education.

### Future directions

Future research should refine prompt engineering to better serve patient education. One potential direction is the integration of automated prompt optimization tools, as demonstrated by Wu JH et al.^25^ Additionally, further studies should assess the effectiveness of formats such as ‘SMART’ in improving models’ accuracy.^37^

Although researchers have tested prompt engineering in ophthalmology and surgery, more work is needed to see how well it applies across other healthcare areas.

Finally, collaboration between AI developers, healthcare providers, and regulatory bodies will ensure that prompt-engineered LLM outputs align with ethical and clinical best practices.^19,40^

### Limitations of This Review

This systematic review has several limitations. The small number of included studies limits generalizability of findings. Heterogeneity in study tasks and methodologies prevents conducting a meta-analysis.

Additionally, rapid evolution of LLM technology means that the performance reported in existing studies may not accurately represent the models’ current or future capabilities.

## Conclusions

This review shows that well-designed prompt engineering often improves readability and clarity in LLM-generated patient education. Accuracy gains vary across models and fields. Studies reveal a trade-off between precision and accessibility. More trials are needed to define standard methods, control bias, and refine evaluations. These steps can yield reliable, accessible patient information.

## Data Availability

All data produced in the present study are available upon reasonable request to the authors

## Data availability statement

All data analyzed during this study are publicly accessible online.

## Ethics Approval Statement

Ethical approval is not applicable for this article.

## Patient Consent Statement

There are no human participants in this article and informed consent is not required.

## Author Contribution

All authors contributed to the study conception. The methodology design and material preparation were performed by AM, EK, and SS. Data collection, formal analysis, writing of the original draft, and figure creation were conducted by AM. Validation, supervision, and project administration were handled by EK and SS. All authors commented on previous versions of the manuscript. All authors have read and approved the final manuscript.

## REFERENCES

1. Bhattad PB, Pacifico L. Empowering Patients: Promoting Patient Education and Health Literacy. Cureus. 2022;14(7):e27336. doi:10.7759/cureus.27336

2. Stacey D, Légaré F, Lewis K, et al. Decision aids for people facing health treatment or screening decisions. Cochrane Database Syst Rev. 2017;4(4):CD001431. doi:10.1002/14651858.CD001431.pub5

3. Dineen-Griffin S, Garcia-Cardenas V, Williams K, Benrimoj SI. Helping patients help themselves: A systematic review of self-management support strategies in primary health care practice. PLoS One. 2019;14(8):e0220116. doi:10.1371/journal.pone.0220116

4. Stiller C, Brandt L, Adams M, Gura N. Improving the Readability of Patient Education Materials in Physical Therapy. Cureus. 2024;16(2):e54525. doi:10.7759/cureus.54525

5. Bedi S, Jain SS, Shah NH. Evaluating the clinical benefits of LLMs. Nat Med. 2024;30(9):2409–2410. doi:10.1038/s41591-024-03181-6

6. Aydin S, Karabacak M, Vlachos V, Margetis K. Large language models in patient education: a scoping review of applications in medicine. Front Med (Lausanne). 2024;11:1477898. doi:10.3389/fmed.2024.1477898

7. Singhal K, Tu T, Gottweis J, et al. Toward expert-level medical question answering with large language models. Nat Med. Published online January 8, 2025. doi:10.1038/s41591-024-03423-7

8. Mudrik A, Nadkarni GN, Efros O, Glicksberg BS, Klang E, Soffer S. Exploring the role of Large Language Models in haematology: A focused review of applications, benefits and limitations. Br J Haematol. Published online September 3, 2024. doi:10.1111/bjh.19738

9. Meskó B. Prompt Engineering as an Important Emerging Skill for Medical Professionals: Tutorial. J Med Internet Res. 2023;25:e50638. doi:10.2196/50638

10. Amisha, Malik P, Pathania M, Rathaur VK. Overview of artificial intelligence in medicine. J Family Med Prim Care. 2019;8(7):2328–2331. doi:10.4103/jfmpc.jfmpc_440_19

11. Ting Sim JZ, Fong QW, Huang W, Tan CH. Machine learning in medicine: what clinicians should know. Singapore Med J. 2023;64(2):91–97. doi:10.11622/smedj.2021054

12. Joshi AK. Natural language processing. Science. 1991;253(5025):1242-1249. doi:10.1126/science.253.5025.1242

13. Soffer S, Ben-Cohen A, Shimon O, Amitai MM, Greenspan H, Klang E. Convolutional Neural Networks for Radiologic Images: A Radiologist’s Guide. Radiology. 2019;290(3):590–606. doi:10.1148/radiol.2018180547

14. Sorin V, Barash Y, Konen E, Klang E. Deep Learning for Natural Language Processing in Radiology-Fundamentals and a Systematic Review. J Am Coll Radiol. 2020;17(5):639–648. doi:10.1016/j.jacr.2019.12.026

15. Ashish Vaswani, Noam Shazeer, Niki Parmar, Jakob Uszkoreit, Llion Jones, Aidan N. Gomez, Łukasz Kaiser, and Illia Polosukhin. 2017. Attention is all you need. In Proceedings of the 31st International Conference on Neural Information Processing Systems (NIPS’17). Curran Associates Inc., Red Hook, NY, USA, 6000–6010.

16. Almarie B, Teixeira PEP, Pacheco-Barrios K, Rossetti CA, Fregni F. Editorial - The Use of Large Language Models in Science: Opportunities and Challenges. Princ Pract Clin Res. 2023;9(1):1–4. doi:10.21801/ppcrj.2023.91.1

17. Fink A, Rau A, Kotter E, Bamberg F, Russe MF. [Optimized interaction with Large Language Models[]: A practical guide to Prompt Engineering and Retrieval-Augmented Generation]. Radiologie (Heidelberg, Germany). Published online February 21, 2025. doi:10.1007/s00117-025-01416-2

18. Zaghir J, Naguib M, Bjelogrlic M, Névéol A, Tannier X, Lovis C. Prompt Engineering Paradigms for Medical Applications: Scoping Review. J Med Internet Res. 2024;26:e60501. doi:10.2196/60501

19. Shah K, Xu AY, Sharma Y, et al. Large Language Model Prompting Techniques for Advancement in Clinical Medicine. J Clin Med. 2024;13(17). doi:10.3390/jcm13175101

20. Chan CM, Xu C, Yuan R, et al. RQ-RAG: Learning to Refine Queries for Retrieval Augmented Generation. Published online March 31, 2024.

21. Roelle J, Müller C, Roelle D, Berthold K. Learning from instructional explanations: effects of prompts based on the active-constructive-interactive framework. PLoS One. 2015;10(4):e0124115. doi:10.1371/journal.pone.0124115

22. Zirui Song, Bin Yan, Yuhan Liu, et al. Injecting Domain-Specific Knowledge into Large Language Models: A Comprehensive Survey. Feb 2025. 10.48550/arXiv.2502.10708.

23. Badarudeen S, Sabharwal S. Assessing readability of patient education materials: current role in orthopaedics. Clin Orthop Relat Res. 2010;468(10):2572–2580. doi:10.1007/s11999-010-1380-y

24. Readable. (n.d.). Automated Readability Index. https://readable.com/readability/automated-readability-index/.

25. Wu JH, Nishida T, Moghimi S, Weinreb RN. Effects of prompt engineering on large language model performance in response to questions on common ophthalmic conditions. Taiwan J Ophthalmol. 2024;14(3):454–457. doi:10.4103/tjo.TJO-D-23-00193

26. Readable. (n.d.). Flesch Reading Ease and Flesch-Kincaid Grade Level. https://readable.com/readability/flesch-reading-ease-flesch-kincaid-grade-level/.

27. Jung H, Oh J, Stephenson KAJ, Joe AW, Mammo ZN. Prompt engineering with ChatGPT3.5 and GPT4 to improve patient education on retinal diseases. Can J Ophthalmol. Published online September 5, 2024. doi:10.1016/j.jcjo.2024.08.010

28. Kianian R, Sun D, Crowell EL, Tsui E. The Use of Large Language Models to Generate Education Materials about Uveitis. Ophthalmol Retina. 2024;8(2):195–201. doi:10.1016/j.oret.2023.09.008

29. Ellison IE, Oslock WM, Abdullah A, et al. De novo generation of colorectal patient educational materials using large language models: Prompt engineering key to improved readability. Surgery. 2025;180:109024. doi:10.1016/j.surg.2024.109024

30. Readable. (n.d.). SMOG Index. https://readable.com/readability/smog-index/.

31. Readable. (n.d.). Gunning Fog Index. https://readable.com/readability/gunning-fog-index/.

32. Readable. (n.d.). Coleman-Liau Readability Index. https://readable.com/readability/coleman-liau-readability-index/.

33. Moher D. Preferred Reporting Items for Systematic Reviews and Meta-Analyses: The PRISMA Statement. Ann Intern Med. 2009;151(4):264. doi:10.7326/0003-4819-151-4-200908180-00135

34. McInnes MDF, Moher D, Thombs BD, et al. Preferred Reporting Items for a Systematic Review and Meta-analysis of Diagnostic Test Accuracy Studies. JAMA. 2018;319(4):388. doi:10.1001/jama.2017.19163

35. Moons KGM, de Groot JAH, Bouwmeester W, et al. Critical Appraisal and Data Extraction for Systematic Reviews of Prediction Modelling Studies: The CHARMS Checklist. PLoS Med. 2014;11(10):e1001744. doi:10.1371/journal.pmed.1001744

36. Whiting PF. QUADAS-2: A Revised Tool for the Quality Assessment of Diagnostic Accuracy Studies. Ann Intern Med. 2011;155(8):529. doi:10.7326/0003-4819-155-8-201110180-00009

37. Vaira LA, Lechien JR, Abbate V, et al. Enhancing AI Chatbot Responses in Health Care: The SMART Prompt Structure in Head and Neck Surgery. OTO Open. 2025;9(1):e70075. doi:10.1002/oto2.70075

38. Ahn S. The impending impacts of large language models on medical education. Korean J Med Educ. 2023;35(1):103–107. doi:10.3946/kjme.2023.253

39. Lee D, Palmer E. Prompt engineering in higher education: a systematic review to help inform curricula. International Journal of Educational Technology in Higher Education. 2025;22(1):7. doi:10.1186/s41239-025-00503-7

40. Goktas P, Grzybowski A. Shaping the Future of Healthcare: Ethical Clinical Challenges and Pathways to Trustworthy AI. J Clin Med. 2025;14(5):1605. doi:10.3390/jcm14051605

