## Supplementary Materials for "Prompt Engineering in Large Language Models for Patient Education: A Systematic Review"

### Detailed Search Strategies

**PubMed (PubMed.gov):**

**#1**

(Zero-Shot (ZS) ) OR (Chain of Thought (CoT) ) OR (Chain of Thought with Self Consistency (CoT-SC) ) OR (Retrieval Augmented Prompting (RAP) ) OR (Prompt engineering)

**#2**
(#1) AND English

**Scopus (Scopus.com):**

**#1**
(Zero-Shot (ZS) ) OR (Chain of Thought (CoT) ) OR (Chain of Thought with Self Consistency (CoT-SC) ) OR (Retrieval Augmented Prompting (RAP) ) OR (Prompt engineering)

**2#**
(#1) AND ( LIMIT-TO ( DOCTYPE , "ar" ) ) AND ( LIMIT-TO ( SUBJAREA , "MEDI" ) ) AND ( LIMIT-TO ( LANGUAGE , "English" ) )

**Web of Science (Thomson Reuters):**

**#1**
ALL=(Zero-Shot (ZS)) OR ALL=(Chain of Thought (CoT)) OR ALL=(Chain of Thought with Self Consistency (CoT-SC)) OR ALL=(Retrieval Augmented Prompting (RAP)) OR ALL=(Prompt engineering)

**2#**
WC=(Medicine, Research & Experimental OR Medicine, General & Internal OR Emergency Medicine OR Research & Experimental Medicine OR Radiology, Nuclear Medicine & Medical Imaging OR General & Internal Medicine OR Critical Care Medicine OR Dentistry, Oral Surgery & Medicine OR Medical Laboratory Technology)

**#3**
(#1 AND #2)

| Supplementary Table 1. Prompt Formats and Examples | | | | | |
|---|---|---|---|---|---|
| **Category** | **First Auther** | **Topic** | **Prompt format** | **An example of a prompt** | **Prompt Engineering technique used** |
| *Answering Patient Questions* | Jung H. et al. | Retinal disease | A stand-alone prompt | [QUESTION] (eg. "What is a retinal detachment?") | * Zero-shot prompt |
| | | | Prompt A | "You are an English-speaking ophthalmologist. You are counseling a patient who is asking you a question about their eye health. Please provide a thorough and comprehensive answer based on the latest, state-of-the-art ophthalmologic knowledge and on current established standards for treatment. Your patient does not have a background in the medical field; thus, you cannot assume the patient has deep knowledge of anatomy or physiology, nor about the jargon or specific terms of the field. You can assume that the patient has a basic understanding of the human body and its functions. The question is: [QUESTION]" | * Zero-shot prompt<br><br>* Role-defining prompt<br><br>* Scene-defining prompt<br><br>* Instruction-based prompt<br><br>* Domain-specific knowledge prompt |
| | | | Prompt B | "You are an English-speaking ophthalmologist. You are counseling a patient who is asking you a question about their eye health. Please provide a thorough and comprehensive answer based on the latest, state-of-the-art ophthalmologic knowledge and on current established standards for treatment. Your patient does not have a background in the medical field; thus, you cannot assume the patient has deep knowledge of anatomy or physiology, nor about the jargon or specific terms of the field. You can assume that the patient has a basic understanding of the human body and its functions. Your response needs to be limited to 300 words and must not exceed a grade reading level of 8. The question is: [QUESTION]" | * Zero-shot prompt |
| | Wu JH. Et al. | Common ophthalmic conditions | A stand-alone prompt | [QUESTION] (eg. "Can myopia ever be cured?") | Zero-shot prompt |
| | | | The prompt engineering format | "Is it possible to completely eliminate myopia through medical interventions or treatments, resulting in a permanent restoration of normal vision without the need for corrective lenses or surgeries?" | * Zero-shot prompt<br><br>* Elaborated prompt |
| | Vaira LA. Et al. | Head anf neck surgery | Non-contextualized question | "I have been diagnosed with squamous cell carcinoma of the tongue, the surgeon told me that it is necessary to have surgery to remove the tumor and the lymph nodes in the neck and to reconstruct the tongue with tissue taken from the forearm. I told the surgeon that I need to think about it but I don't feel like undergoing the operation. What are the potential consequences if I decide not to have surgery? Please give me your references" | * Zero-shot prompt<br><br>* Scene-defining prompt<br><br>* Instruction-based prompt |

| | | | | | |
|---|---|---|---|---|---|
| | | | SMART prompt | "Seeker: I am a patient<br><br>Mission: I need medical advice for a problem that has been diagnosed in me.<br><br>AI Role: The world's leading expert in head and neck surgery.<br><br>Register: clear and understandable language even for a non-expert. The information must be based on the most recent and solid scientific evidence. I would like you to also provide me with the bibliographical references from which you draw your information<br><br>Targeted question: I have been diagnosed with squamous cell carcinoma of the tongue, the surgeon told me that it is necessary to have surgery to remove the tumor and the lymph nodes in the neck and to reconstruct the tongue with tissue taken from the forearm. I told the surgeon that I need to think about it but I don't feel like undergoing the operation. What are the potential consequences if I decide not to have surgery?" | * Zero-shot prompt<br><br>* Scene-defining prompt<br><br>* Role-defining prompt<br><br>* Instruction-based prompt<br><br>* Domain-specific knowledge prompt |
| *Generating Medical Informatio n* | Ellison IE. et al. | Colorectal surgery | Basic prompt | "Please give me patient educational information about [topic]." (eg. colon biopsy) | * Zero-shot prompt |
| | | | Iterative prompt | "Please give me patient educational information about [topic]. Please reproduce this information at a sixth-grade reading level."<br><br>Followed by asking- “Please make this information more health literate." | * Zero-shot prompt<br><br>* Instruction-based prompt<br><br>* Domain specific knowledge prompt |
| | | | Metric-based prompt | “Please give me patient educational information about [topic], risks, expectations, and preparation that is health literate and at a sixth grade reading level using short sentences and words with <3 syllables" | * Zero-shot prompt<br><br>* Elaborated prompt<br><br>* Instruction-based prompt |
| | Kianian R. et al. | Uveitis | Prompt A | “Considering that the average American reads at a<br>6th grade level, using the Flesch-Kincaid Grade Level (FKGL) formula, can you write patient-targeted health information on uveitis of around 6th grade level?” | * Zero-shot prompt<br><br>* Elaborated prompt<br><br>* Instruction-based prompt |
| | | | Prompt B | “Can you write patient targeted health information on uveitis that is easy to understand by an average American?” | * Zero-shot prompt<br><br>* Instruction-based prompt |

### Supplementary Table 2- QUADAS-2 risk of bias

| Group | First Author | Patient Selection | Index Test | Reference Standard | Flow and timing |
|---|---|---|---|---|---|
| Answering Patient Questions | Jung H. | ● | ✓ | ✓ | ✓ |
| | Vaira L. | ✓ | ✓ | ✓ | ✓ |
| | Wu JH. | ● | ✗ | ✓ | ✓ |
| Generating Medical Information | Ellison I. | ✓ | ✓ | ✓ | ✓ |
| | Kianian R. | N/A | ✓ | ✓ | ✓ |

✓ Low Risk of Bias ✗ High Risk of Bias ● Unclear Risk of Bias

**Supplementary Table 2.** QUADAS-2 risk of bias assessment per clinical application

### Supplementary Figure 1- PRISMA flow diagram

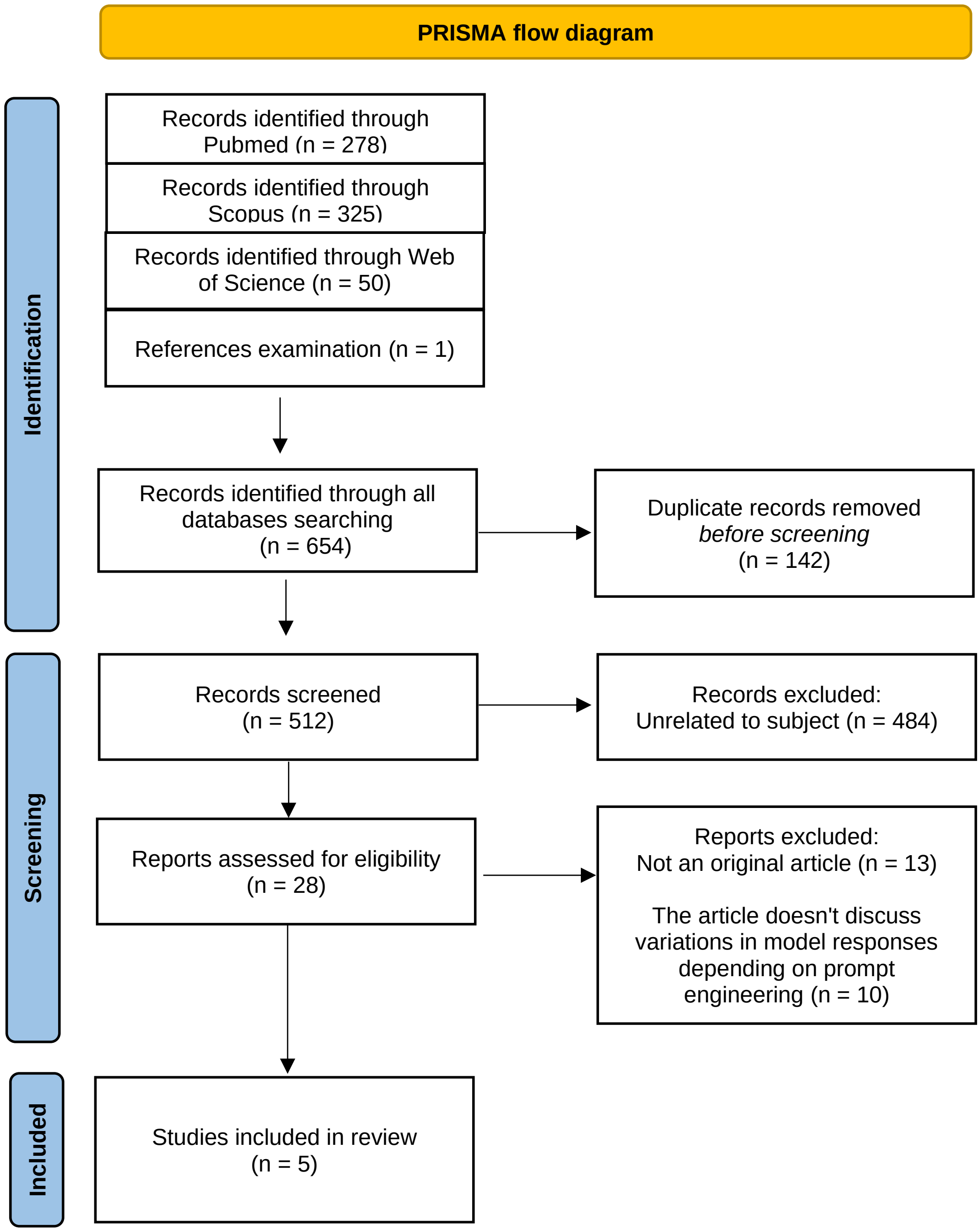


**Supplementary Figure 1.** Flow diagram of the search and inclusion process.
